# Pharmacokinetic analysis of carboplatin and fluorescein brain permanence following ultrasound-based blood-brain barrier opening

**DOI:** 10.1101/2025.01.20.25320847

**Authors:** Karl J. Habashy, Timothy W. Synold, Ye Feng, Cristal Gomez, Christina Amidei, Rachel Ward, Sarah VanderMolen, Ashkan Zarrieneh, Kwang-Soo Kim, Victor A. Arrieta, Jawad Fares, Kirsten B. Burdett, Hui Zhang, Crismita Dmello, Li Chen, John F. Bebawy, Michael Canney, Roger Stupp, Behnam Badie, Jana Portnow, Adam M. Sonabend

**Affiliations:** Department of Neurological Surgery, Feinberg School of Medicine, Northwestern University, Chicago, IL, USA; Northwestern Medicine Lou & Jean Malnati Brain Tumor Institute of the Lurie Comprehensive Cancer Center, Feinberg School of Medicine, Northwestern University, Chicago, IL, USA; Analytical Pharmacology Core, Beckman Research Institute, City of Hope Comprehensive Cancer Center, Duarte, CA, USA; Department of Preventive Medicine, Feinberg School of Medicine, Northwestern University, Chicago, IL, USA; Department of Anesthesiology, Feinberg School of Medicine, Northwestern University, Chicago, IL, USA; Carthera, Lyon, France; Department of Neurology, Feinberg School of Medicine, Northwestern University, Chicago, IL, USA; Division of Hematology/Oncology, Department of Medicine, Feinberg School of Medicine, Northwestern University, Chicago, IL, USA; Department of Neurosurgery, City of Hope Comprehensive Cancer Center, Duarte, CA, USA; Department of Medical Oncology, City of Hope Comprehensive Cancer Center, Duarte, CA, USA

## Abstract

**Background:** The blood-brain barrier (BBB) impedes the passage of most circulating drugs into the brain. Low-intensity pulsed ultrasound with microbubbles (LIPU/MB) transiently opens the BBB, improving parenchymal drug penetration. Parenchymal drug permanence upon short-lived BBB opening is unknown. We compared the parenchymal permanence of temozolomide, carboplatin, and fluorescein, and investigated the effect of LIPU/MB on the concentration of carboplatin and fluorescein.

**Methods:** We analyzed four patients who underwent intraoperative LIPU/MB with intravenous administration of carboplatin and fluorescein in the NCT04528680 clinical trial. Microdialysis catheters were implanted into sonicated and non-sonicated brain and measured drug levels over 24 hours. Published data from a microdialysis study of temozolomide without LIPU/MB were used for comparison.

**Results:** LIPU/MB led to sustained elevated parenchymal drug concentrations, achieving 3.1-fold increase in brain-to-plasma AUC for carboplatin and fluorescein (P = 0.03). Compared to non-sonicated brain, sonicated brain had higher concentrations of carboplatin for 11 hours, and fluorescein for 5 hours. Drug levels in the sonicated brain exceeded their plasma concentrations at 21 hours and 7 hours, for carboplatin and fluorescein, respectively. In non-sonicated brain, drug half-life was longest for fluorescein (13.6 ± 11.0 hours), followed by carboplatin (5.1 ± 1.9 hours) and temozolomide (2.9 ± 1.6 hours). Sonication did not affect parenchymal drug half-life.

**Conclusion:** Following LIPU/MB, BBB-impermeable drugs exhibit sustained elevated parenchymal concentrations surpassing their plasma levels, highlighting the bi-directional restriction of drug passage by the BBB. Future studies are warranted to explore drug trapping and the efficacy of sustained exposure to cytotoxic drugs for the treatment of brain-infiltrating tumors.

## Introduction

The blood-brain barrier (BBB) is a semi-permeable structure that limits the passage of most drugs from the systemic circulation into the brain.^1,2^ Several characteristics influence drug permeability across the BBB, such as molecular weight, lipophilicity, polar surface area, hydrogen bonding capacity, and the amount of unbound drug in the plasma.^3,4^ In addition, efflux transporters such as multidrug-resistant P-glycoproteins, predominantly expressed on the luminal side of endothelial cells, transport penetrant hydrophobic drugs back into the blood.^5–7^ In this context, extensive efforts have been dedicated to characterizing the optimal drug formulations for improved delivery, with an emphasis on the design of hydrophobic drugs with low molecular weight.

Yet, while the role of the BBB in limiting drug penetration into the brain is extensively described, it is possible that this barrier also limits the leakage of agents from the human brain parenchyma back into the circulation. Drugs with limited BBB permeability and resistance to efflux by multidrug-resistant proteins may therefore have prolonged retention in the brain compared to BBB-permeable drugs, which are often substrates for efflux pumps.^5–7^ While the downstream implications on drug bioavailability and permanence in the brain include potential effects on therapeutic efficacy and duration of drug action, human data supporting of drug trapping in the brain is scarce.

Low-intensity pulsed ultrasound with concomitant administration of microbubbles (LIPU/MB) transiently opens the BBB and improves the delivery of cytotoxic drugs and antibodies by multiple folds.^8–16^ Following LIPU/MB, the BBB in humans is predominantly restored within an hour.^8,11^ It is not known whether drugs that enter into the brain at the time of LIPU/MB-based BBB opening, can be retained in the parenchyma once the BBB integrity is restored following LIPU/MB.

In this study, we examined the concentration of carboplatin and fluorescein over 24 hours in the plasma and brain regions that were subject to LIPU/MB shortly before intravenous carboplatin administration. Parenchymal concentrations in sonicated and non-sonicated brain were measured using intracerebral microdialysis catheters, which capture free-drug (unbound) levels capable of exerting a therapeutic effect. In the context of intact BBB (no sonication), we also compared the pharmacokinetics of carboplatin and fluorescein, compounds with limited BBB permeability,^8,17^ to those of temozolomide (TMZ), a relatively BBB-permeable agent with pharmacokinetic data previously reported by Portnow J, et al.^18,19^

## Results

### In vitro fractional recovery

The average fractional recovery of carboplatin at 0.5 µL/min was 65% ± 9% (average ± sd) and was similar across both tested concentrations (P > 0.05; Supplementary Figure 1). Increasing the flow rate to 1 µL/min or 2 µL/min decreased the fractional recovery to 38% and 23%, respectively (P < 0.01). A flow rate of 0.5 µL/min was therefore adopted for *in vivo* studies. In that context, we evaluated the fractional recovery of fluorescein at that flow rate and recorded an average recovery of 41% ± 10% (Supplementary Figure 1).

### Study participants

A total of 6 patients were selected for this pharmacokinetic study, including two females and four males, with ages ranging from 48 to 72 years. Intraoperative sonication involved the activation of 3 ultrasound emitters over the peritumoral normal brain (Figure 1A). Each participant had a catheter implanted into a sonicated brain region, identified by fluorescein extravasation, and another catheter implanted into non-sonicated brain (Figure 1B). Catheter placement in normal non-enhancing brain was confirmed using intraoperative neuronavigation and further corroborated post-operatively with fused MRI and computer tomography (CT) scan (Figure 1C). Two patients were excluded due to technical difficulties with catheter placement. We therefore had paired drug level measurements from sonicated and non-sonicated brain in 4 patients (8 catheters).

**Figure 1.**
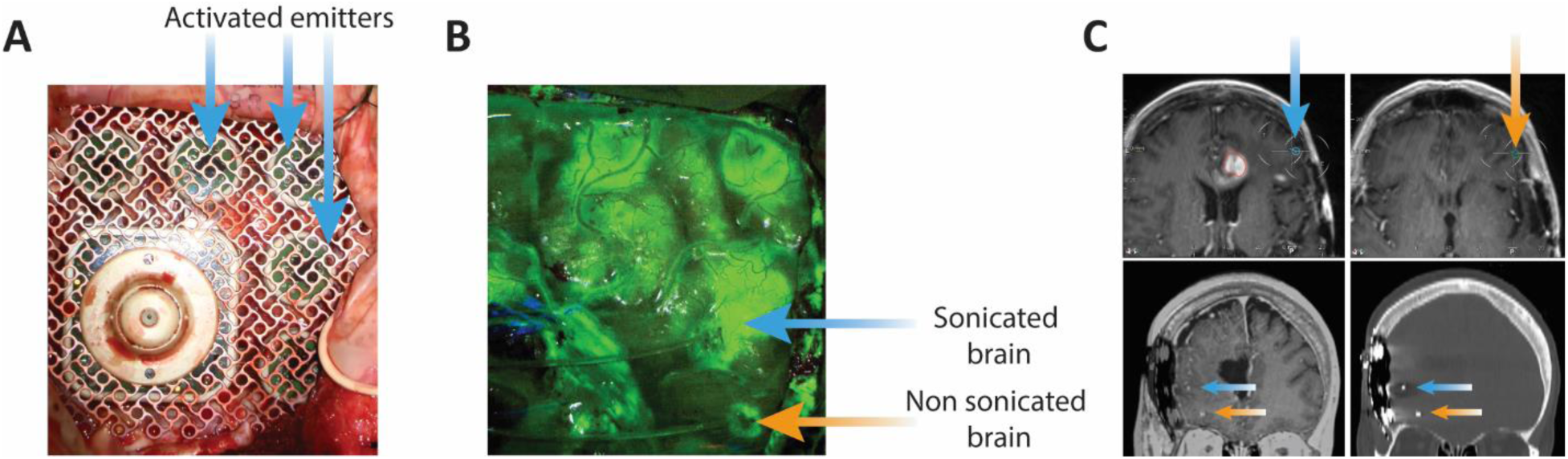
Intraoperative procedure with LIPU/MB and microdialysis implantation. (A) Surgical microscope images illustrating the placement of the SonoCloud-9 ultrasound device over a skull window for intraoperative sonication with microbubbles. The blue arrows designate the ultrasound emitters that were activated. (B) Fluorescent microscope image illustrating the extravasation of fluorescein into the sonicated brain. The blue arrow points to the insertion site of the first microdialysis catheter into sonicated fluorescent brain. The orange arrow points to the insertion site of the second microdialysis catheter into non-fluorescent, non-sonicated brain. (C) Intraoperative navigation MRI illustrating the placement of the microdialysis catheters in sonicated (blue arrow) and non-sonicated (orange arrow) non-enhancing brain (upper panel). In the lower panel, post-operative MRI fused with CT scan (left) and CT scan (right) confirm the placement of the microdialysis catheters into sonicated (blue arrow) and non-sonicated (orange arrow) brain.

### Brain pharmacokinetics of carboplatin and fluorescein with LIPU/MB

We previously demonstrated that most of the BBB is restored within an hour following LIPU/MB, (Supplementary Figure 2) and that LIPU/MB increases the concentration of carboplatin in the peritumoral brain by 5.9-times at approximately this timeframe.^8^ Here, we investigated the free-drug concentration of carboplatin (Figure 2A) and fluorescein (Figure 2B) in the sonicated and non-sonicated brain over 24 hours. Parenchymal drug measurement was initiated 2 hours after sonication, a timepoint where the BBB was predominantly restored. Our first microdialysate was collected 4 hours after LIPU/MB and averaged the parenchymal drug concentration for the period spanning from 2 to 4 hours after LIPU/MB (noted as 3 hours). Subsequent samples were collected at 2 hours intervals.

**Figure 2.**
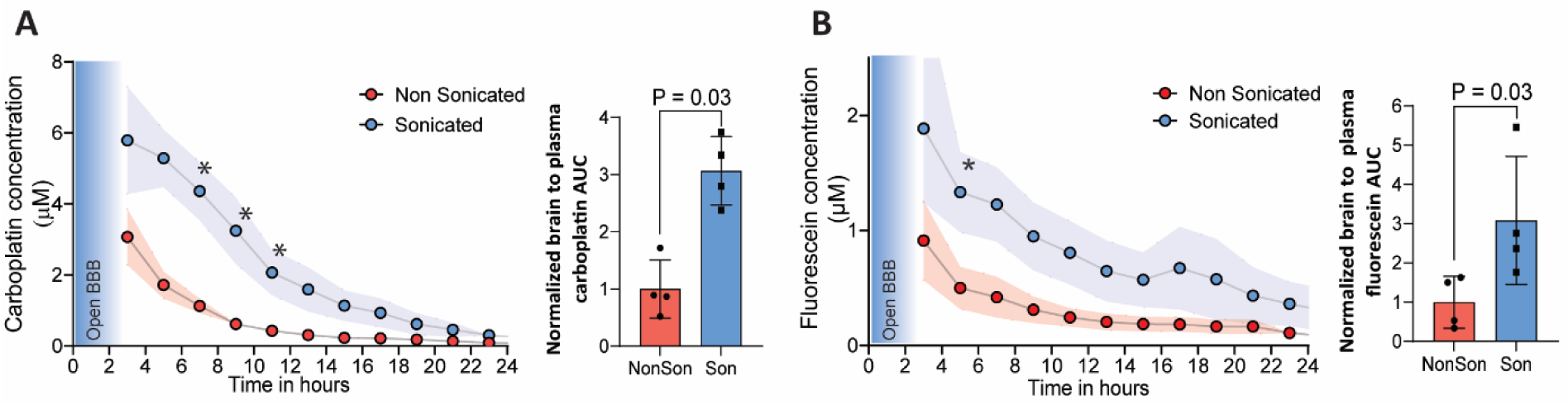
Low-intensity pulsed ultrasound with microbubbles leads to sustained elevation in parenchymal drug levels. Graphs illustrating the concentration over time curves of carboplatin (A) and fluorescein (B) in the sonicated (blue) and non-sonicated (red) brain. The first microdialysate collection measured parenchymal drug levels from 2 to 4 hours after sonication, averaged 3 hours, timepoint where most of the BBB had been restored. The shaded area shows the standard error of the mean. Drug concentrations were higher in sonicated compared to non-sonicated brain up to 11 hours for carboplatin and 5 hours for fluorescein and (P < 0.05). Brain-to-plasma drug concentration over time curves were 3.1 times higher in sonicated compared to non-sonicated brain for both carboplatin and fluorescein (P = 0.03). *, P < 0.05 compared to non-sonicated brain. BBB = Blood-brain barrier; NonSon = Non sonicated; Son = Sonicated.

The average maximal drug concentration (Cmax) reached in the non-sonicated brain was 3.1 μM (95% confidence interval [CI] 0.6 – 5.6 μM) for carboplatin and 0.9 μM (95% CI 0 – 2.0 μM) for fluorescein. In sonicated brain, carboplatin reached a Cmax of 6.4 μM (95% CI 2.9 – 10.0 μM) and fluorescein a Cmax of 1.9 μM (95% CI 0 – 3.9 μM). Individual concentrations can be found in Table 1. In the plasma, the total drug levels (bound and unbound) were measured, revealing a carboplatin Cmax of 55.2 μM (95% CI 21.2 - 89 μM), and a fluorescein Cmax of 134.1 μM (95% CI 100.5 – 167.7 μM).

**Table 1:** Table illustrating the maximal concentration measured in sonicated and non-sonicated brain for carboplatin and fluorescein.

|  | Carboplatin |  | Fluorescein |  |
| --- | --- | --- | --- | --- |
|  | Non-Sonicated Brain | Sonicated Brain | Non-Sonicated Brain | Sonicated Brain |
| **Patient 1** | 4.7 μM | 9.1 μM | 1.5 μM | 3.5 μM |
| **Patient 2** | 1.3 μM | 6.8 μM | 0.2 μM | 1.7 μM |
| **Patient 3** | 4.0 μM | 6.2 μM | 1.5 μM | 2.0 μM |
| **Patient 4** | 2.2 μM | 3.7 μM | 0.4 μM | 0.5 μM |
| **Average** | 3.1 ± 1.6 μM | 6.4 ± 2.2 μM | 0.9 ± 0.7 μM | 1.9 ± 1.2 μM |

Over time, drug concentrations in sonicated brain were persistently higher than in non-sonicated brain. For instance, carboplatin levels in sonicated brain remained higher than in non-sonicated brain for up to 11 hours (P < 0.05; Figure 2A), while fluorescein remained higher for up to 5 hours (P < 0.05; Figure 2B). In the absence of sonication, both drugs had limited permeation into the BBB, with a brain-to-plasma area under the curve (AUC) of 16.3% (95% CI 3.1 – 29.4%) for carboplatin (Figure 2A) and 10.4% (95% CI 0 – 21.3%) for fluorescein (Figure 2B). Sonication enhanced drug delivery to the brain, leading to a 3.1-fold increase in the brain-to-plasma AUC for both carboplatin (AUC 47.9%, 95% CI 14.1 – 81.7%; P = 0.03; Figure 2A) and fluorescein (AUC 27.9%, 95% CI 2.3 – 53.6%; P = 0.03; Figure 2B), highlighting a sustained elevation in parenchymal drug levels with LIPU/MB.

We then investigated whether LIPU/MB-mediated BBB-opening influences drug clearance from the brain. Notably, parenchymal drug half-life was similar in the sonicated and non-sonicated brain, both for carboplatin (mean 5.1 hours [95% CI 2.1 - 8.1 hours] in non-sonicated brain and 4.8 hours [95% CI 2.6 – 7.0 hours] in sonicated brain, P = 0.86) and fluorescein (mean 13.6 hours [95% CI 0 – 31.1 hours] in non-sonicated brain and 7.7 hours [95% CI 3.0 – 12.5 hours] in sonicated brain, P = 0.13). Sonication therefore had negligeable effect on parenchymal drug clearance, likely due to the rapid closure of the BBB following LIPU/MB (Supplementary Figure 3). It is likely that following LIPU/MB, parenchymal drug levels and permanence are influenced by Cmax, which can be enhanced by LIPU/MB, and by the drug’s parenchymal half-life, which is not affected by LIPU/MB.

### Permanence of chemotherapeutic drugs in the brain

Considering that LIPU/MB did not affect parenchymal drug half-life, and that prolonged drug permanence in the brain may be necessary for adequate therapeutic effect, we sought to investigate whether the chemical properties of drugs influence their clearance from the brain. We therefore hypothesized that BBB-impermeable drugs may have prolonged brain permanence due to their limited diffusion across the intact BBB in both directions. Since brain pharmacokinetic data with LIPU/MB is scarce, we compared the clearance of carboplatin and fluorescein from non-sonicated brain, to that of temozolomide (TMZ), previously published by Portnow and colleagues,^18^ as that study was performed using the same protocol and technique. Indeed, similarly to our study, TMZ’s brain concentration was measured using microdialysis catheters implanted in non-enhancing normal brain.^18^ Importantly, TMZ, carboplatin, and fluorescein share similar molecular weights but variable characteristics such as lipophilicity, polar surface area, and hydrogen bonding capacity influencing their BBB permeability (Figures 3A, B, C). TMZ is a relatively BBB-permeable drug,^18,19^ while carboplatin and fluorescein have limited penetration of the BBB.^8,17^

**Figure 3.**
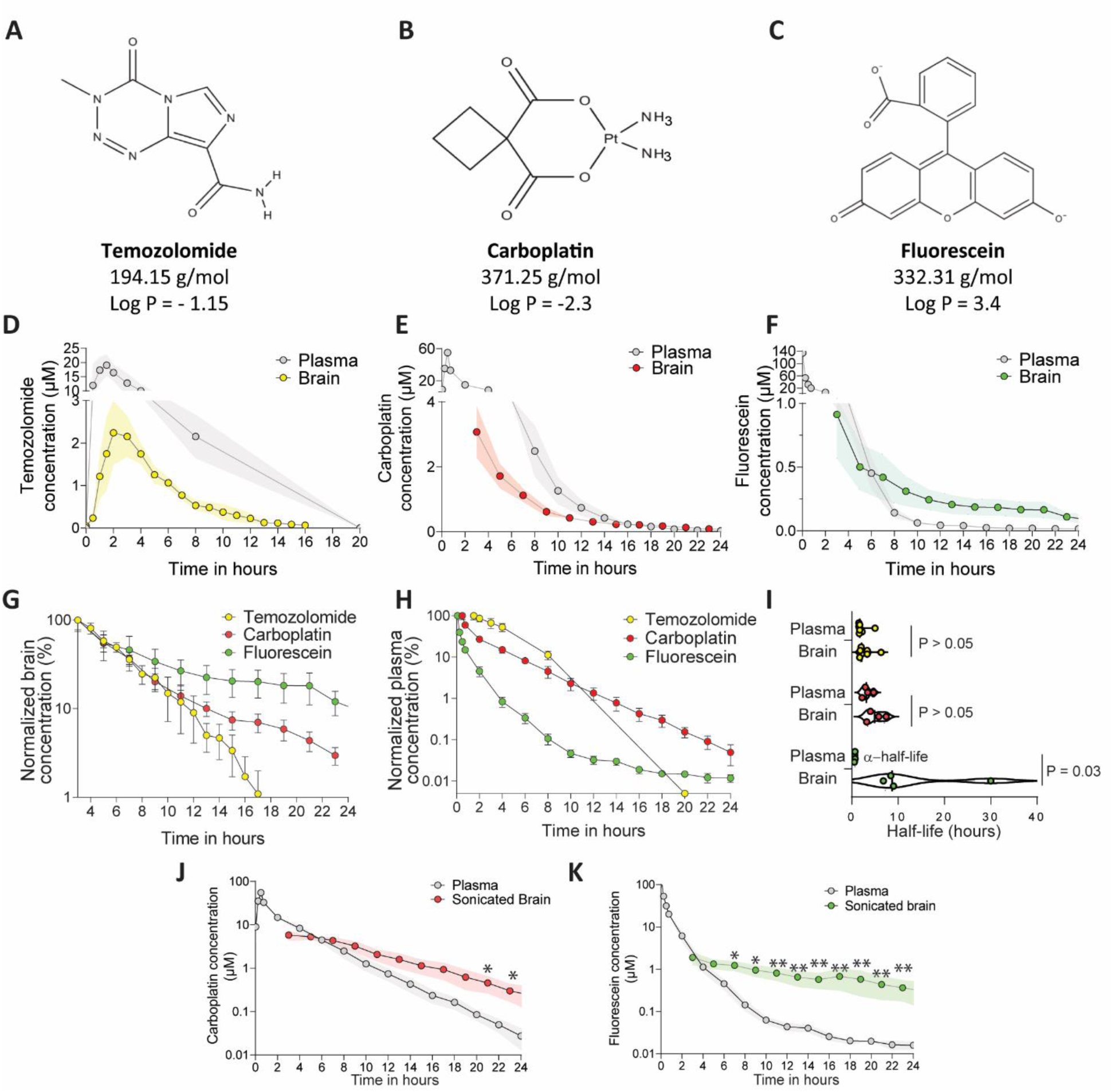
Brain parenchymal permanence of temozolomide, carboplatin, and fluorescein. (A, B, C) Chemical structure, molecular weight, and partition coefficient of temozolomide (A), carboplatin (B), and fluorescein (C) as reported on the National Institute of Health’s National Center for Biotechnology Information and on Screnci D, et al., Br J of Cancer, 2000. (D, E, F) Drug concentration over time curves illustrating the plasma and brain parenchymal concentrations of temozolomide (D), carboplatin (E), and fluorescein (F) in normal non-sonicated brain. (G, H) Graphs illustrating the rate of decrease of drug levels from the brain (G) and plasma (H). Drug concentrations are normalized to their highest concentrations and shown on a logarithmic scale. Within the brain, temozolomide has the shortest half-life followed by carboplatin and fluorescein. In plasma, fluorescein concentrations decreased the fastest due to its short initial half-life, followed by temozolomide and carboplatin. (I) Violin plot illustrating the plasma and parenchymal half-life of temozolomide, carboplatin, and fluorescein. We illustrate here the initial half-life of fluorescein as the drug followed a two-phase decay. The half-life of temozolomide and carboplatin in brain and plasma are similar, while that of fluorescein is longer in the brain (P = 0.03). (J, K), Drug concentration over time curves illustrating that the concentration of carboplatin (J) and fluorescein (K) in sonicated brain (free-drug) exceed those in the plasma (free and bound drug). *, P < 0.05; **, P < 0.01.

Drug clearance from the brain interstitial space was fastest for TMZ which had an average parenchymal half-life of 2.9 hours (95% CI 1.4 - 4.4 hours) and was nearly undetected at 16 hours post-infusion (Figures 3D, G). Carboplatin had a slightly longer parenchymal half-life of 5.1 hours (95% CI 2.1 - 8.1 hours, P = 0.35 vs TMZ; Figures 3E, G), while drug permanence in the brain was the longest for fluorescein that reached a parenchymal half-life of 13.6 hours (95% CI 0 – 31.1 hours, P = 0.005 vs TMZ; Figures 3F, G). In contrast, drug clearance from the plasma was fastest for fluorescein (initial half-life 0.6 hours, 95% CI 0.5 - 0.7 hours; Figures 3F, H), followed by TMZ (half-life of 2.1 hours, 95% CI 1.1 - 3.1 hours, P = 0.07 vs fluorescein; Figures 3D, H) and carboplatin (half-life of 3.3 hours, 95% CI 1.6 - 5.0 hours, P = 0.004 vs fluorescein; Figure 3E, H). Fluorescein’s plasma concentration exhibited a two-phase decay with a terminal half-life reaching 9.6 hours (95% CI 2.3 - 16.9 hours). However, 77% (95% CI 71 – 82%) of its plasma concentration was cleared within 30 minutes of the bolus infusion. The plasma half-lives of TMZ and carboplatin were similar to their brain parenchymal half-lives in (P > 0.05). However, the parenchymal half-life of fluorescein was larger than its plasma initial half-life (P = 0.03; Figure 3I), illustrating likely delayed clearance from the brain compared to plasma. Our results support the hypothesis that BBB-permeable drugs are cleared from the brain as they are metabolized systemically, while BBB-impermeable drugs may become trapped within the brain parenchyma.

In support of this, we observed that the parenchymal free-drug concentrations of TMZ remained consistently below its plasma concentration (Figure 3D), while the parenchymal concentrations of carboplatin and fluorescein reached levels comparable to those of total (bound and unbound) drug in the plasma (Figures 3E, F). However, as plasma measurements included bound and free-drug levels, the true difference between parenchymal and plasma free-drug concentrations may be underestimated. It is therefore likely that parenchymal free-drug levels of carboplatin and fluorescein in non-sonicated brain exceeded their corresponding plasma free-drug levels. In the context of sonication, the parenchymal free-drug levels exceeded plasma total drug levels starting at 21 hours for carboplatin (Figure 3H) and 7 hours for fluorescein (Figure 3K), highlighting the trapping of BBB-impermeable drugs in the brain parenchyma.

## Discussion

Three pathways have been suggested to be involved in drug clearance from the brain: 1) transport across the BBB back into circulation, 2) transport through the cerebrospinal fluid or glymphatic system, and 3) local metabolism into metabolites that are further eliminated through routes 1 and/or 2.^20^ Little is known about the influence of drug properties on their permanence in the human brain parenchyma and elimination rate. Here, we illustrated that the permanence of BBB-permeable drugs such as TMZ in the human brain is determined by their plasma half-life, confirming observations reported in preclinical models, including with other chemotherapies such as busulfan.^4,21,22^ It is therefore likely that, as these drugs are metabolized and cleared systemically, they cross back from the brain parenchyma into systemic circulation, reaching an equilibrium, and are rapidly cleared from the brain.

In contrast, we observed that fluorescein, which is less BBB-permeable than TMZ, accumulated within the brain parenchyma where the levels of free drug surpassed the plasma concentration of total (protein-bound and free) drug as early as 7 hours following LIPU/MB, and up to 24 hours. Unlike BBB-permeable drugs that are cleared from the brain by diffusing freely across the BBB, BBB-impermeable drugs may become trapped and have a delayed clearance from the parenchyma. In support of this, a previous intracerebral microdialysis study in patients with recurrent glioma found that methotrexate exhibited prolonged brain permanence in non-enhancing regions with intact BBB compared to enhancing regions with disrupted BBB.^23^ Consistent findings have been reported in preclinical models where doxorubicin accumulated in the brain following improved delivery with ultrasound, while its concentration decreased over 24 hours in tumor tissue with disrupted BBB.^24^ Moreover, a preclinical microdialysis study illustrated a prolonged brain permanence of morphine 6-glucuronide (M6G) compared to its more BBB-permeable predecessor, morphine, despite both having a similar systemic half-life.^25^ It is therefore likely that the BBB acts as a two-way restrictive barrier trapping impermeable drugs within the brain parenchyma and slowing their clearance through transport into systemic circulation. Indeed, while studies have primarily focused on the role of the BBB in limiting influx of molecules into the brain, the restrictive properties also apply to the efflux of molecules, as suggested by the presence of transporters to clear brain metabolites with limited BBB permeability.^20,26^ This two-way property of the BBB is also evidenced by the release of cell free nucleic acids from the brain following LIPU/MB, as we and others have reported.^27–29^

In this context, a fast restoration of the BBB following LIPU/MB is crucial to trap brain-impermeable drugs in the brain parenchyma following their enhanced delivery during the short window of BBB opening. We recently described that most of the BBB is repaired within an hour following LIPU/MB,^8,11^ which is faster than the timeframe previously reported in preclinical and clinical studies,^30–32^ stressing the importance of this period for BBB homeostasis and restoration after injury. In a single-cell transcriptomic and ultrastructural analysis of sonicated and non-sonicated human brain samples collected within this critical timeframe, we observed distinct endothelial cell gene expression changes after LIPU/MB, particularly in genes related to neurovascular barrier function and structure, including changes to genes involved in the basement membrane, endothelial cell cytoskeleton, and junction complexes, as well as caveolar transcytosis and various solute transporters.^33^ Ultrastructural analysis of capillary endothelium showed that LIPU/MB led to a decrease in luminal caveolae, the emergence of vacuoles, and the disruption of the basement membrane and tight junctions. ^33^

Thus, it is likely that the active process of BBB homeostasis and restoration following LIPU/MB results in the decrease of the leakage of drugs from the brain parenchyma, and explains the retention of carboplatin and fluorescein in the sonicated brain at concentrations exceeding their plasma levels. Supporting the concept of BBB-impermeable drug trapping in the brain following transient LIPU/MB-based BBB opening, we reported in a separate study of patients with glioblastoma a 2-fold increase in the concentration of doxorubicin and pembrolizumab, two relatively BBB-impermeable drugs, in peri-tumoral sonicated brain biopsies two days after sonication.^15^

Our analyses and results focus on the human brain, in contrast to other pharmacokinetic studies performed within the tumor where the BBB is disrupted.^23,34^ This is especially important in the context of glioma treatment since lesions that exhibit disrupted BBB and Gadolinium enhancement are typically resected, leaving infiltrative glioma cells in non-enhancing peritumoral areas with intact BBB. The penetration of drugs specifically into these peri-tumoral non-enhancing region is key for treating the infiltrative disease.

Our results suggest that LIPU/MB may better combine with BBB-impermeable drugs due to an initial increase in parenchymal drug concentrations during the open BBB window (first hour), followed by a delayed drug clearance and prolonged permanence of unbound active drug in the brain upon BBB restoration.

Our study has several limitations. First, intracerebral microdialysis relies on sensitive membranes that can be disrupted during placement or with patient movement, potentially affecting drug measurements. In addition, catheter migration, occlusion or kinking can compromise the ability to perform the pharmacokinetic study. Second, we compared a limited number of drugs and patients who met demanding clinical and anatomical criteria that were conducive for this intraoperative and perioperative complex pharmacokinetic study. Third, due to the integration of intraoperative LIPU/MB into the neurosurgical routine for tumor resection, we were unable to obtain samples within the first 1-2 hours after sonication. Finally, it is also important to note that we report here the free-drug concentrations in the brain, while plasma concentrations represent total (free and bound) drug levels. It is therefore likely that the true extent of drug trapping in the brain is underestimated in this study.

## Conclusion

The human BBB appears to act as a bi-directional restrictive membrane limiting the diffusion of BBB-impermeable drugs from parenchyma back into systemic circulation. Combining LIPU/MB with impermeable drugs might be advantageous due to prolonged parenchymal drug permanence, at elevated, biologically meaningful concentrations.

## Materials and Methods

### Study participants and design

A convenience subset of patients from the non-randomized, multi-center phase 1/2 clinical trial (NCT04528680) for patients with recurrent glioblastoma was enrolled when microdialysis implant and intraoperative sonication were possible. Patients on the trial were above 18 years of age, had a confirmed diagnosis of isocitrate dehydrogenase 1 (IDH1) wild-type glioblastoma with radiographic evidence of recurrence or progression after failure of 1 or 2 lines of therapy, and a maximal tumor diameter below 70 mm on magnetic resonance imaging (MRI) prior to surgery for resection of the recurrent tumor. Patients selected for the microdialysis pharmacokinetic study included those who did not require corticosteroids during the surgery or perioperative time and had tumors located in non-eloquent brain regions, with a tumor cavity smaller than the ultrasound device, enabling sonication and microdialysis sampling of peri-tumoral sonicated and non-sonicated brain. Patients were enrolled from a single tertiary care center from 2023 to 2024.

### Intraoperative procedure

Patients received surgery for resection of the recurrent tumor and implantation of a 9-emitter ultrasound device (SonoCloud-9, Carthera, Lyon, France) as described on our phase 1 trial.^8^ Intraoperative sonication was performed with the activation of 3 of 9 ultrasound emitters over the peritumoral brain with an intravenous administration of microbubbles (Definity 10 μL/kg, Lantheus), followed by intravenous infusion of carboplatin (over 30 minutes, at an area under the curve [AUC] of 5) and fluorescein (500 mg intravenous bolus). Fluorescein, which does not readily cross the intact BBB, was injected within minutes of sonication for identification of sonicated areas with disrupted BBB.^8^ Microdialysis catheters (70 Brain MD catheter 100/10, M Dialysis) were then implanted into peritumoral sonicated and non-sonicated brain for drug sampling over 24 hours. Microdialysis catheter placement was guided by MRI neuronavigation and fluorescent microscopy for placement in non-enhancing sonicated and non-sonicated normal brain. Catheters were perfused with central nervous system (CNS) perfusion fluid (Perfusion Fluid CNS, M Dialysis) and microdialysate samples were collected every 2 hours for a total of 24 hours. Due to the integration of this pharmacokinetic study into the neurosurgical routine for tumor resection, resection of the tumor was performed first after sonication, and drug sampling with microdialysis was initiated 2 hours post-sonication, with the first sample collected 4 hours post-sonication. For every patient, we collected microdialyate samples from sonicated and non-sonicated brain. Plasma samples were collected every 15 minutes for the initial 45 minutes then simultaneously with microdialysate samples. No corticosteroids were administered during surgery or perioperative time. Around two weeks after surgery, patients received carboplatin (AUC = 5) and nab-paclitaxel (80 mg/kg) with LIPU/MB in cycles of three weeks. The Institutional Review Board (IRB) of Northwestern University gave ethical approval for this work in accordance with the institutional ethical regulations and the Declaration of Helsinki principles. Written informed consent was obtained from patients before participation in the trial and pharmacokinetic study. Published data on the peritumoral brain concentration of TMZ for 7 patients were used for comparison.^18^

### In vitro determination of fractional recovery

An *in vitro* assessment of fractional recovery was performed at drug concentrations observed in the human brain as reported in Sonabend AM, et al.^8^ Briefly, microdialysis catheters were placed into a reservoir containing carboplatin (2 µM or 10 µM) or fluorescein (10 µM), and microdialysate was collected in triplicates at flow rates of 0.5 µL/min, 1 µL/min, and 2 µL/min. The fractional recovery was used to correct for the *in vivo* results.

### Liquid chromatography tandem mass spectrometry

Plasma samples underwent protein precipitation using acetonitrile, followed by centrifugation at 15,000 × g for 5 min at 4°C to yield clear supernatants. These supernatants were subsequently diluted and injected into the LC-MS/MS system. Specifically, 50 μL of each plasma sample, along with 5.0 μL of an internal standard (IS) was mixed with 200 μL of acetonitrile. After centrifugation, the supernatant was further diluted with either organic or aqueous solvent: for carboplatin, 50 μL of supernatant was mixed with 150 μL of acetonitrile; for fluorescein, 50 μL of supernatant was mixed with 150 μL of water with 0.1% formic acid.

Dialysate samples were analyzed using a dilute and shoot strategy. Specifically, 50 μL of each dialysate sample was first mixed with the appropriate IS. For carboplatin analysis, 10 μL of this mixture was then diluted with 200 μL of acetonitrile; for fluorescein, 10 μL of the mixture was mixed with 200 μL of a 20% acetonitrile solution with 0.1% formic acid.

Quantification of carboplatin and fluorescein levels was conducted using a Shimadzu Prominence HPLC system coupled with an AB SCIEX QTRAP® 5500 mass spectrometer. The column temperature was maintained at 40°C, and the auto-injector temperature was set at 5°C.

Chromatographic separation for carboplatin utilized a Kinetex HILIC analytical column (2.6 µm, 50 × 2.0 mm) with a Phenomenex HILIC guard column, operating at a flow rate of 0.2 mL/min. The mobile phase consisted of solvent A (0.1% acetic acid in 90% acetonitrile) and solvent B (water with 0.1% formic acid). The gradient program spanned from 0% to 95% B over 1 to 2.6 minutes, held at 95% B for 2.6 to 5 minutes, returned to 0% B over 5 to 5.1 minutes, and maintained at 0% B from 5.1 to 7 minutes. The total run time was 7 minutes, with retention times for carboplatin and its internal standard (Carboplatin-D4) at 1.60 minutes.

For fluorescein, a Luna C8 column (2.6 µm, 50 × 2.0 mm) was employed at a flow rate of 0.3 mL/min. The mobile phase comprised solvent A (water with 0.1% formic acid) and solvent B (acetonitrile). Gradient elution ranged from 20% to 50% B over 0 to 1.5 minutes, 50% to 100% B over 1.5 to 2 minutes, 100% B over 2 to 4 minutes, and back to 20% B over 4 to 5 minutes. Retention times for fluorescein and its internal standard Zen3694 were 1.45 and 2.25 minutes, respectively.

The electrospray ionization source operated in positive ion mode with optimized parameters for ion source gases, ionspray voltage, source temperature, declustering potential (DP), and collision energy (CE). Multiple reaction monitoring (MRM) was employed for quantitation, with precursor/product ion transitions as follows: m/z 372.0→294.0 for carboplatin, 377.0→299.0 for carboplatin-D4, 333.1→287.2 for fluorescein, and 334.2→243.2 for Zen3694. Data acquisition and processing were conducted using Analyst software version 1.6.1.

### Pharmacokinetic analysis

Concentration over time curves were plotted using the measured plasma and corrected microdialysate concentrations for each patient using non-compartmental methods. The AUC was calculated using the rule of linear trapezoids extrapolated to infinity. Half-lives (t_1/2_) were calculated from the elimination rate constant derived from the last six measured concentrations. Fluorescein concentration in the brain exhibited a two-phase decay, the initial half-life was derived from the initial four measured concentrations. In comparing brain-to-plasma drug levels, microdialysate (collected over 2 hours) concentrations were compared to the average plasma drug concentration from samples collected at the start and end of the 2-hour microdialysate collection periods.

### Statistical analysis

A one-way analysis of variance (ANOVA) test was used to compare the *in vitro* data that were normally distributed as confirmed by the Shapiro-Wilk test. The remaining statistical analysis of *in vivo* data was performed using non-parametric tests to account for the limited sample size. The Mann-Whitney test was used for two-samples comparisons. The Kruskal-Wallis and Friedman tests with Dunn’s multiple comparison were performed for unpaired and paired comparisons, respectively. P values below 0.05 were considered statistically significant and Dunn’s adjustment was applied when necessary. Statistical analysis was performed using GraphPad (Prism v.9).

## Funding

National Institutes of Health (1R01CA245969-01A1) to A.M.S & R.S, (P50CA221747) to A.M.S. & R.S., (1U19CA264338-01) to A.M.S. and R.S., (U19CA264512) to J.P., B.B, and T.S. A.M.S also received funding support from the Lou and Jean Malnati Brain Tumor Institute, philanthropic support from the Moceri Family Foundation as well as Tina and Vic Kedaitis.

## Disclosures

AMS and RS have received research support from Carthera, BMS and Agenus. AMS is a paid consultant for Carthera and EnClear Therapeutics. RS serves as a member of the scientific advisory board of Alpheus Medical, Carthera, and has provided consultancy to AstraZeneca, Boston Scientific, Novartis and Novocure. RS has equity in Alpheus Medical and Carthera.MC is a full-time employee of Carthera and holding equity. Remaining authors: none declared.

## Data Availability

Data included in this study is available upon request.

---

**Supplementary Figure 1.**
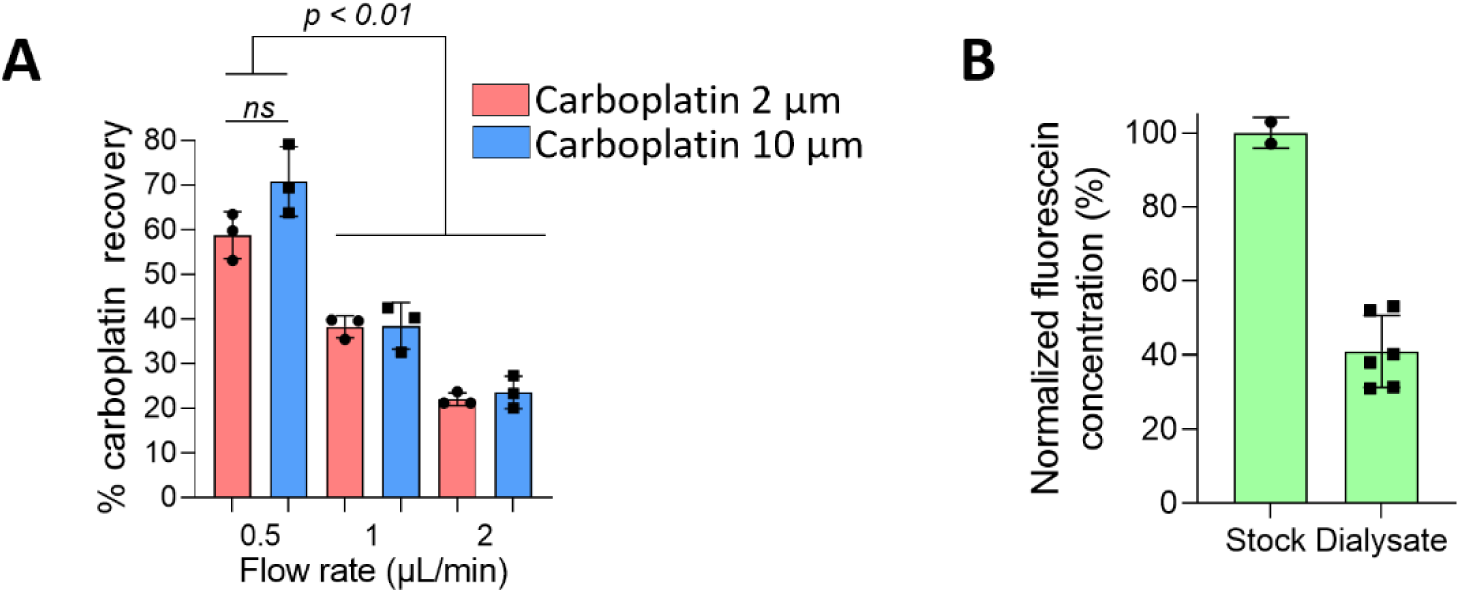
In vitro fractional recovery of carboplatin and fluorescein (A) Graph illustrating the *in vitro* fractional recovery of carboplatin at concentrations of 2 μM and 10 μM, and flow rates of 0.5 μL/min, 1 μL/min, and 2 μL/min. The highest recovery was recorded at 0.5 μL/min for both concentrations (P < 0.01). This flow rate was then considered for fluorescein (B), which resulted in a recovery of 41%.

**Supplementary Figure 2.**
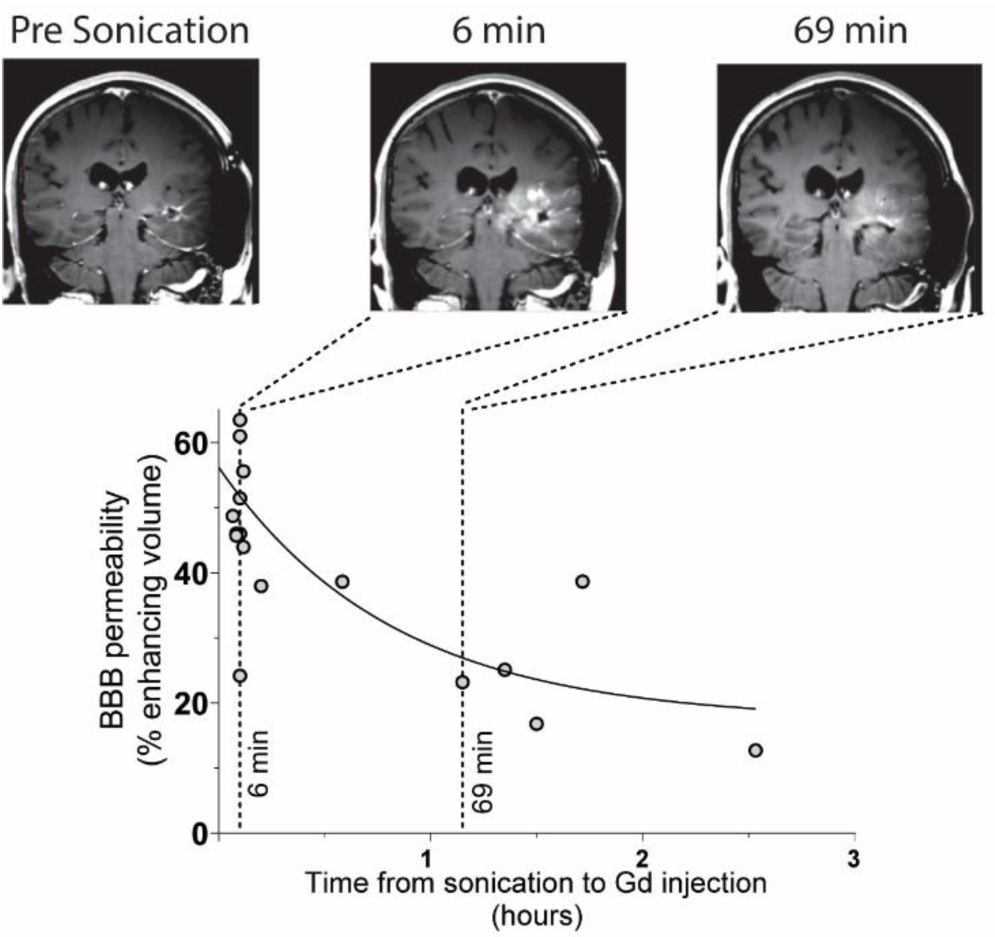
The blood brain barrier is restored promptly following low-intensity pulsed ultrasound and microbubbles. MRI-based analysis of BBB repair following LIPU/MB, adapted from Sonabend et al., Lancet Onc, 2023. The study comprised an analysis of Gadolinium enhancement during 19 sonication cycles in 17 patients, correlating the level of enhancement with the time from sonication to contrast injection. The study demonstrates that most of the BBB is repaired in the first hour following LIPU/MB.

**Supplementary Figure 3.**
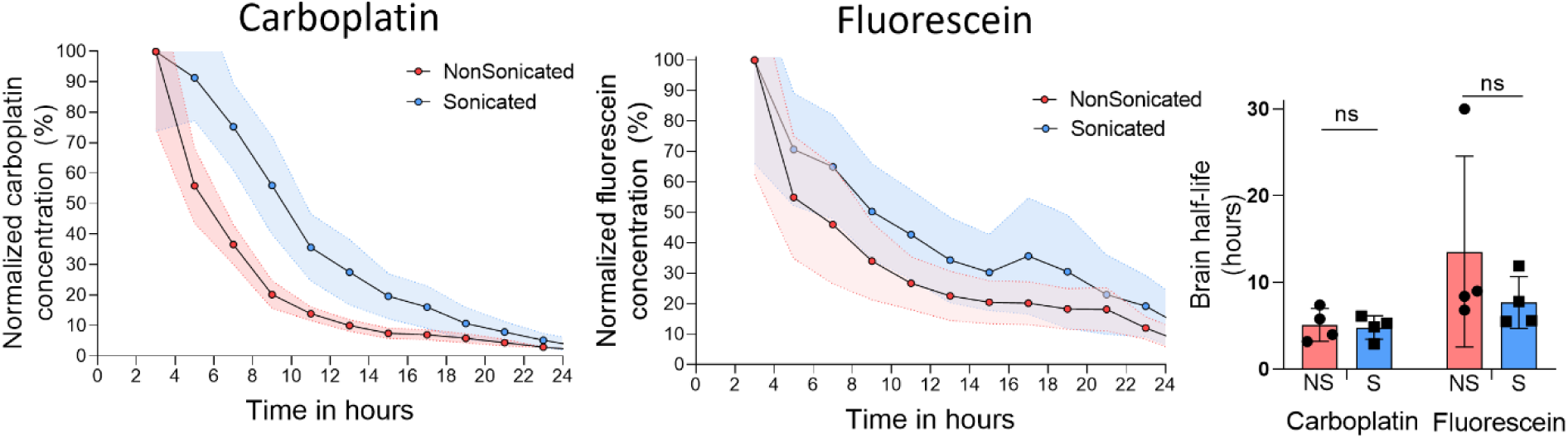
Sonication does not affect drug half-life in the brain parenchyma. Graphs illustrating the rate of carboplatin and fluorescein clearance from the sonicated (blue) and non-sonicated (red) brain parenchyma. We show on the right the half-life quantification of drug permanence in the brain, illustrating no differences between sonicated and non-sonicated brain for both drugs. NS = Non Sonicated; S = Sonicated.

## Notes

### Author Declarations

The Institutional Review Board (IRB) of Northwestern University gave ethical approval for this work.

